# Detection of Severe Structural Heart Disease Using an AI-Enhanced Portable 1-Lead ECG: The ACCESS-SHD Study

**DOI:** 10.64898/2026.09.14.26361683

**Authors:** Arya Aminorroaya, Sumukh Vasisht Shankar, Mariam Khan, Madeleine Carter, Lovedeep S Dhingra, Akshay Khunte, Bernardo Lombo, Robert L McNamara, Evangelos K Oikonomou, Aline F Pedroso, Rohan Khera

**Affiliations:** Section of Cardiovascular Medicine, Department of Internal Medicine, Yale School of Medicine, New Haven, CT, USA; Cardiovascular Data Science (CarDS) Lab, Yale School of Medicine, New Haven, CT, USA; NYU Grossman School of Medicine, New York, NY, USA; Section of Health Informatics, Department of Biostatistics, Yale School of Public Health, New Haven, CT, USA; Department of Biomedical Informatics and Data Science, Yale School of Medicine, New Haven, CT, USA; Center for Outcomes Research and Evaluation, Yale-New Haven Hospital, New Haven, CT, USA

**Keywords:** **Keywords**: Portable Electrocardiography, KardiaMobile, Structural Heart Disease, Deep Learning, Screening

## Abstract

**Aims:** Structural heart disease (SHD) often remains undetected until symptoms develop. Portable 1-lead ECG devices return an automated rhythm-based interpretation, but whether AI-ECG adds diagnostic value beyond this interpretation is unknown. We prospectively evaluated a noise-adapted 1-lead AI-ECG algorithm for detecting severe SHD from portable KardiaMobile 6L recordings, and its relative diagnostic value beyond the device’s rhythm interpretation.

**Methods and Results:** Adults undergoing outpatient echocardiography between June 2024 and January 2025 recorded a 30-second, 1-lead ECG with real-time AI-ECG inference. The primary endpoint was discrimination for echocardiography-defined severe SHD. Secondary analyses assessed net reclassification improvement (NRI) versus the native interpretation and the number needed to test (NNT). Among 597 participants (median age 61.7 years; 51.4% women), 30 (5.1%) had severe SHD. AI-ECG achieved an AUROC of 0.872 (95% CI: 0.806– 0.938), meeting the prespecified endpoint, with 86.7% (70.3–94.7) sensitivity, 72.5% (68.7– 76.0) specificity, 99.0% (97.5–99.6) negative predictive value, and 14.4% (10.1–20.3) positive predictive value, with comparable performance across subgroups. AI-ECG increased sensitivity by 34.6 percentage points (95% CI: 13.0–56.0) over the native interpretation and yielded a categorical NRI of 24.3% (95% CI: 2.8–45.9), with 76.9% sensitivity and 80.0% specificity among tracings the device read as normal. An AI-ECG-guided strategy for detecting severe SHD reduced the NNT from 19.7 to 6.9 (a 64.8% reduction).

**Conclusions:** A noise-adapted AI-ECG algorithm detected severe SHD from real-world portable 1-lead ECGs and substantially improved case finding beyond the device’s rhythm interpretation, supporting AI-ECG-guided triage as a potential scalable screening strategy.

## INTRODUCTION

Structural heart disease (SHD) is frequently recognized only after symptoms or complications develop, despite the availability of effective therapies for phenotypes such as left ventricular systolic dysfunction (LVSD) and left-sided valvular disease.^1–7^ Community echocardiographic screening has shown that a large share of SHD remains undiagnosed in the population, particularly among older adults.^6–8^ Because earlier identification could enable timely treatment, when therapeutic benefit is greatest, scalable strategies to detect SHD before it becomes clinically apparent are needed.^5–7^ Yet transthoracic echocardiography (TTE), the reference standard for diagnosis, is too resource-intensive to deploy at the population scale.^9–11^ The electrocardiogram (ECG), by contrast, is inexpensive, ubiquitous, and increasingly obtainable outside healthcare settings, making it an attractive substrate for scalable screening.^12–14^

Artificial intelligence-enhanced ECG (AI-ECG) has been developed to detect SHD from both 12- and 1-lead recordings, on the premise that structural abnormalities produce electrical signatures not discernible to human readers.^15–18^ Although 12-lead AI-ECG can identify a range of SHDs, its role in community-based screening is limited; 1-lead ECG, in contrast, is uniquely suited to screening outside clinical settings through portable, consumer-grade devices.^19,20^ AI-ECG models applied to lead I of clinical ECGs, signals that approximate those captured by portable devices, can detect a composite of clinically actionable SHD,^15^ yet validation on recordings from actual portable 1-lead devices has largely been confined to isolated phenotypes such as LVSD, whose individually low prevalence constrains positive predictive value in screening.^21–24^ Furthermore, enthusiasm for the capacity of AI-ECG to detect SHD has outpaced evidence on whether it adds value beyond the interpretation already available to users. Portable 1-lead devices return a rhythm-based interpretation directly to the consumer;^25–27^ whether, and to what extent, AI-ECG improves SHD detection beyond this native interpretation has, to our knowledge, not been examined. These devices are particularly useful as, unlike wearable consumer devices with 1-lead ECG capabilities that are restricted to a single user, the portable devices can be used to acquire 1-lead ECGs in a community setting from a large number of individuals.

In the ACCESS-SHD study, we prospectively evaluated a noise-adapted 1-lead AI-ECG algorithm, developed and validated previously,^15^ for detecting severe SHD from 1-lead ECGs acquired with the portable KardiaMobile 6L device among patients undergoing TTE. We aimed to assess the algorithm’s standalone diagnostic performance and to quantify its incremental value over the device’s native rhythm-based interpretation.

## METHODS

### Study Design

ACCESS-SHD was a prospective diagnostic validation study conducted in the echocardiography laboratory of Yale New Haven Hospital (YNHH). Adults presenting for a clinically indicated TTE were invited to record a 1-lead ECG using a portable KardiaMobile 6L device during the same visit, after which the tracings were linked to structured TTE reports and electronic health record (EHR) data using unique participant identifiers (**Figure 1**). The study was designed to evaluate, at the point of care, both the standalone accuracy of an AI-ECG algorithm for detecting severe SHD and its diagnostic value relative to the portable device’s own native interpretation. The study protocol was approved by the Yale Institutional Review Board (#2000035532). The data from the Yale New Haven Health System represent protected health information. To protect patient privacy, the Yale Institutional Review Board does not allow the sharing of these data.

**Figure 1.**
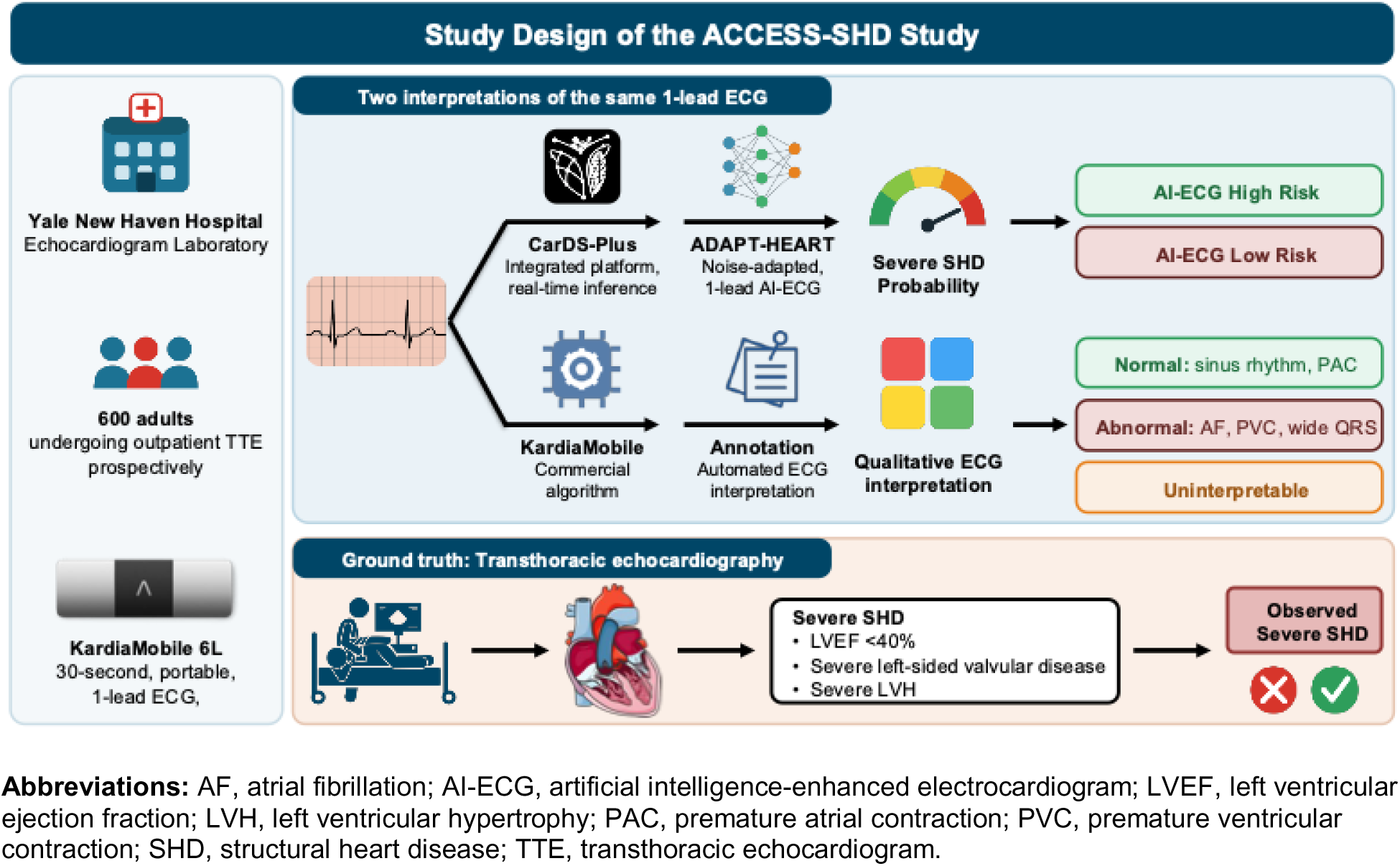
Study Design of the ACCESS-SHD Study.

### Study Population

We prospectively enrolled adults aged 18 years or older who were scheduled for an outpatient TTE at the YNHH echocardiography laboratory between June 2024 and January 2025. Individuals with a permanent pacemaker or implantable cardioverter-defibrillator, those who were pregnant, those with cognitive impairment, and those unable to communicate in English were not eligible. All participants provided written informed consent before enrollment.

### Study Covariates

Severe SHD, the primary target condition, was defined as a composite of any of the following findings on TTE: a left ventricular ejection fraction (LVEF) below 40%, denoting LVSD; severe aortic or mitral stenosis or regurgitation; or severe left ventricular hypertrophy (LVH), defined as an interventricular septal thickness in diastole greater than 15 mm accompanied by moderate or severe left ventricular diastolic dysfunction. We additionally defined a broader SHD composite that incorporated moderate or severe, rather than only severe, left-sided valvular disease. Echocardiographic measurements adhered to the recommendations of the American Society of Echocardiography.^28^ LVEF was quantified using the Simpson biplane method, or by three-dimensional echocardiography or visual estimation when biplane imaging was unavailable, and valvular and diastolic function were graded by the interpreting cardiologist.^29^

Clinical covariates, comprising hypertension, type 2 diabetes, atherosclerotic cardiovascular disease (coronary artery disease, ischemic stroke, transient ischemic attack, or peripheral artery disease), and heart failure, were ascertained from the EHR using International Classification of Diseases, Tenth Revision, Clinical Modification codes, as described previously.^30–32^

### 1-lead ECG Acquisition

After providing consent, each participant recorded a single 30-second, 1-lead ECG using the KardiaMobile 6L device (AliveCor) during the TTE visit. Participants rested both hands on the device electrodes, in accordance with the manufacturer’s instructions, to obtain a lead I tracing, which was then stored for analysis.

### AI-ECG Model and Prospective Inference

We deployed ADAPT-HEART, a previously developed and externally validated ensemble deep learning algorithm that detects severe SHD from lead I of the ECG.^15^ The algorithm was originally trained on clinical ECGs and explicitly adapted to tolerate the signal noise characteristic of portable and wearable recordings.^21^ During each study visit, the recorded 1-lead tracing was processed through an automated interface that performed inference in real time.^33^ Following the development pipeline, each 30-second recording was divided into 5 overlapping 10-second segments; the waveforms were resampled from 300 Hz at acquisition to 500 Hz and rescaled to match the development data; the ensemble AI-ECG model was applied to each segment; and the median of the segment-level probabilities was taken as the recording-level output. For each outcome label, the corresponding label-specific model was applied.

### KardiaMobile’s Native Rhythm and Conduction Interpretation

To benchmark AI-ECG against the information available to consumers, we recorded the automated rhythm interpretation returned by the KardiaMobile device for every tracing.^25,34,35^ While the device’s automated interpretation is not intended to detect severe SHD, given it is available to the consumer, it could serve as a benchmark to assess whether AI-ECG provides incremental information for detecting severe SHD. For the head-to-head comparison, native interpretations were dichotomized as normal (sinus rhythm or premature atrial contractions) or abnormal (atrial fibrillation, premature ventricular contractions, or wide QRS complexes).

Tracings that the device was unable to interpret were classified as uninterpretable; these were excluded from analyses comparing AI-ECG with the native interpretation but were retained for analyses of standalone AI-ECG performance. The AI-ECG probability was dichotomized at a prespecified threshold chosen to achieve greater than 90% specificity in the internal validation set of the original development study.

### Study Outcomes

The primary outcome was the discrimination of AI-ECG for TTE-defined severe SHD, quantified by the area under the receiver operating characteristic curve (AUROC). Secondary analyses reported threshold-based sensitivity, specificity, NPV, and PPV for severe SHD, the broader SHD composite, and individual phenotypes. To characterize the added value of AI-ECG over the native interpretation, we examined reclassification and the differences in diagnostic performance between the two approaches. Screening efficiency was summarized as the number needed to test (NNT) to identify one case under a usual-care strategy and under an AI-ECG-guided strategy.

### Sample Size Calculation

The sample size was determined a priori for the primary endpoint of severe SHD. Assuming an expected AUROC of approximately 0.91 and an event prevalence of approximately 4 to 5% from preliminary data, enrollment of 600 participants was projected to provide 80% power at a two-sided α of 0.05 while ensuring that the lower bound of the 95% confidence interval (CI) for the AUROC exceeded 0.75, after allowing for attrition of 10% owing to technical or acquisition-related failures.

### Statistical Analysis

Continuous variables were summarized as medians with interquartile ranges and categorical variables as counts with percentages. The AUROC and its 95% CI were estimated using the DeLong method, and sensitivity, specificity, NPV, and PPV using Wilson score intervals.^36^ To quantify the incremental value of AI-ECG beyond the native interpretation, we computed the categorical net reclassification improvement (NRI) for severe SHD, reporting its event and non-event components, and compared sensitivity, specificity, PPV, and NPV between the two approaches.^37^ Analyses involving the native interpretation were restricted to participants with an interpretable KardiaMobile tracing, whereas standalone AI-ECG performance was assessed in all analyzable recordings. The NNT under usual care was derived from the outcome prevalence and the NNT under the AI-ECG strategy from the model’s PPV at the prespecified threshold, with the relative reduction reported. Analyses were conducted in Python (version 3.11), with two-sided tests and a significance level of 0.05.

## RESULTS

### Study Population

Of 600 participants undergoing outpatient TTE who were enrolled, 3 were excluded for failure to capture a 1-lead ECG, leaving 597 with an analyzable recording (**Table 1**). The median age was 61.7 years (IQR, 45.9–71.6), 307 (51.4%) were women, and self-reported race and ethnicity were White in 371 (65.3%), Black in 105 (18.5%), and Hispanic in 54 (9.5%). Hypertension was present in 404 (67.7%), type 2 diabetes in 168 (28.1%), atherosclerotic cardiovascular disease in 288 (48.2%), and heart failure in 164 (27.5%). On TTE, 30 participants (5.1%) met criteria for severe SHD and 97 (16.4%) for SHD. Individual phenotypes included LVSD in 15 (2.5%), severe left-sided valvular disease in 6 (1.0%), moderate or severe left-sided valvular disease in 79 (13.4%), and severe LVH in 9 (1.5%). The KardiaMobile native interpretation classified 478 tracings as sinus rhythm, 80 as clinically abnormal, and 26 as atrial fibrillation, whereas 13 were uninterpretable. The prevalence of severe SHD increased across these categories, from 2.8% among sinus-rhythm tracings to 11.4%, 15.4%, and 30.8%, respectively. Notably, of the 13 tracings the device deemed uninterpretable, 4 (30.8%) had severe SHD, and the AI-ECG model nonetheless produced a risk estimate for each of these recordings.

**Table 1.** Baseline Characteristics of the Study Population and Prevalence of Structural Heart Disease Stratified by KardiaMobile Interpretation Category.

| Characteristic* | Overall | KardiaMobile Interpretation |  |  |  |
| --- | --- | --- | --- | --- | --- |
|  |  | Sinus Rhythm | Clinically Abnormal | AF | Uninterpretable |
| **Number** | 597 | 478 | 80 | 26 | 13 |
| **Age (years)** | 61.7 [45.9–71.6] | 59.3 [43.1–69.2] | 68.6 [57.8–74.4] | 76.4 [72.1–80.6] | 68.8 [45.0–76.2] |
| **Female Sex** | 307 (51.4%) | 262 (54.8%) | 32 (40.0%) | 7 (26.9%) | 6 (46.2%) |
| **Race and Ethnicity** |  |  |  |  |  |
| White | 371 (65.3%) | 291 (63.8%) | 57 (75.0%) | 17 (70.8%) | 6 (50.0%) |
| Black | 105 (18.5%) | 83 (18.2%) | 13 (17.1%) | 4 (16.7%) | 5 (41.7%) |
| Hispanic | 54 (9.5%) | 49 (10.7%) | 3 (3.9%) | 2 (8.3%) | 0 |
| Asian | 18 (3.2%) | 16 (3.5%) | 1 (1.3%) | 0 | 1 (8.3%) |
| Native American | 2 (0.4%) | 2 (0.4%) | 0 | 0 | 0 |
| Others | 18 (3.2%) | 15 (3.3%) | 2 (2.6%) | 1 (4.2%) | 0 |
| **Hypertension** | 404 (67.7%) | 304 (63.6%) | 65 (81.2%) | 25 (96.2%) | 10 (76.9%) |
| **T2D** | 168 (28.1%) | 125 (26.2%) | 29 (36.2%) | 11 (42.3%) | 3 (23.1%) |
| **CAD** | 254 (42.5%) | 183 (38.3%) | 48 (60.0%) | 16 (61.5%) | 7 (53.8%) |
| **Ischemic Stroke/TIA** | 105 (17.6%) | 68 (14.2%) | 26 (32.5%) | 9 (34.6%) | 2 (15.4%) |
| **ASCVD** | 288 (48.2%) | 208 (43.5%) | 55 (68.8%) | 18 (69.2%) | 7 (53.8%) |
| **HF** | 164 (27.5%) | 110 (23.0%) | 31 (38.8%) | 15 (57.7%) | 8 (61.5%) |
| **Severe SHD** | 30 (5.1%) | 13 (2.8%) | 9 (11.4%) | 4 (15.4%) | 4 (30.8%) |
| **SHD** | 97 (16.4%) | 62 (13.1%) | 17 (21.5%) | 12 (46.2%) | 6 (46.2%) |
| **LVSD (LVEF <40%)** | 15 (2.5%) | 5 (1.0%) | 5 (6.2%) | 2 (7.7%) | 3 (23.1%) |
| **Severe Valvular Disease** | 6 (1.0%) | 3 (0.6%) | 2 (2.5%) | 0 | 1 (7.7%) |
| **Moderate or Severe Valvular Disease** | 79 (13.4%) | 52 (11.0%) | 11 (13.9%) | 10 (38.5%) | 6 (46.2%) |
| **Severe LVH** | 9 (1.5%) | 5 (1.1%) | 2 (2.5%) | 2 (7.7%) | 0 |

\*Data are presented as median [interquartile range], or number (percentage).
**Abbreviations:** AF, atrial fibrillation; ASCVD, atherosclerotic cardiovascular disease; CAD, coronary artery disease; HF, heart failure; LVEF, left ventricular ejection fraction; LVH, left ventricular hypertrophy; LVSD, left ventricular systolic dysfunction; SHD, structural heart disease; T2D, type 2 diabetes; TIA, transient ischemic attack.

### Performance of AI-ECG

For detecting severe SHD, AI-ECG achieved an AUROC of 0.872 (95% CI, 0.806–0.938), meeting the prespecified primary endpoint, as the lower bound of the CI exceeded 0.75. At the prespecified threshold, sensitivity was 86.7% (95% CI, 70.3–94.7) and specificity 72.5% (95% CI, 68.7–76.0), with an NPV of 99.0% (95% CI, 97.5–99.6) and a PPV of 14.4% (95% CI, 10.1– 20.3) (**Table 2**). Performance was consistent across demographic and clinical subgroups (**Table 3**). Across the broader SHD composite and individual phenotypes, specificity and NPV remained high while other metrics varied by phenotype (**Table 2**). Detection was strongest for LVSD, with a sensitivity of 93.3% and a specificity of 81.1%, whereas sensitivity for severe valvular disease was comparatively low, with only 6 participants with the disease.

**Table 2.** Performance of AI-ECG for Detecting Composite and Individual Structural Heart Diseases.

| Label* | Sensitivity | Specificity | NPV | PPV | Prevalence | NNT without AI-ECG | NNT with AI-ECG | NNT Reduction |
| --- | --- | --- | --- | --- | --- | --- | --- | --- |
| **Severe SHD** | 86.7% (70.3–94.7) | 72.5% (68.7–76.0) | 99.0% (97.5–99.6) | 14.4% (10.1–20.3) | 30 (5.1%) | 19.7 | 6.9 | 64.8% |
| **SHD** | 56.7% (46.8–66.1) | 78.1% (74.2–81.5) | 90.2% (87.0–92.6) | 33.7% (26.9–41.3) | 97 (16.4%) | 6.1 | 3 | 51.3% |
| **LVSD** | 93.3% (70.2–98.8) | 81.1% (77.7–84.0) | 99.8% (98.8–100) | 11.3% (6.8–18.1) | 15 (2.5%) | 39.7 | 8.9 | 77.7% |
| **Severe Valvular Disease** | 33.3% (9.7–70.0) | 90.1% (87.4–92.3) | 99.2% (98.1–99.7) | 3.3% (0.9–11.4) | 6 (1.0%) | 98.5 | 30 | 69.5% |
| **Moderate or Severe Valvular Disease** | 32.9% (23.6–43.9) | 91.8% (89.1–93.9) | 89.9% (87.0–92.2) | 38.2% (27.6–50.1) | 79 (13.4%) | 7.5 | 2.6 | 65.0% |
| **Severe LVH** | 44.4% (18.9–73.3) | 92.1% (89.6–94.0) | 99.1% (97.9–99.6) | 8.0% (3.2–18.8) | 9 (1.5%) | 65.6 | 12.5 | 80.9% |

\*Data are presented as point estimate (95% CI), number (percentage), or percentage.
**Abbreviations:** AI-ECG, artificial intelligence-enhanced interpretation of electrocardiogram; LVH, left ventricular hypertrophy; LVSD, left ventricular systolic dysfunction; NNT, number needed to test; NPV, negative predictive value; PPV, positive predictive value; SHD, structural heart disease.

**Table 3.** Performance Measures of the AI-ECG Algorithm for Detecting Severe SHD Across Key Subgroups.

| Subgroup* | Sensitivity | Specificity | NPV | PPV | Prevalence | NNT Reduction |
| --- | --- | --- | --- | --- | --- | --- |
| **Overall** | 86.7% (70.3–94.7) | 72.5% (68.7–76.0) | 99.0% (97.5–99.6) | 14.4% (10.1–20.3) | 30 (5.1%) | 64.8% |
| **≥65 years** | 100.0% (75.8–100.0) | 56.4% (50.0–62.5) | 100.0% (97.2–100.0) | 10.4% (6.1–17.4) | 12 (4.8%) | 53.6% |
| **<65 years** | 77.8% (54.8–91.0) | 84.3% (79.9–87.8) | 98.6% (96.3–99.4) | 21.5% (13.3–33.0) | 18 (5.3%) | 75.6% |
| **Female** | 91.7% (64.6–98.5) | 82.8% (78.1–86.7) | 99.6% (97.7–99.9) | 18.0% (10.4–29.5) | 12 (4.0%) | 78.0% |
| **Male** | 83.3% (60.8–94.2) | 61.3% (55.4–67.0) | 98.2% (94.9–99.4) | 12.6% (7.8–19.8) | 18 (6.3%) | 50.2% |
| **White** | 100.0% (75.8–100.0) | 66.7% (61.6–71.4) | 100.0% (98.4–100.0) | 9.2% (5.4–15.4) | 12 (3.3%) | 64.5% |
| **Black** | 86.7% (62.1–96.3) | 78.7% (69.0–85.9) | 97.2% (90.4–99.2) | 40.6% (25.5–57.7) | 15 (14.4%) | 64.5% |
| **Hypertension** | 95.8% (79.8–99.3) | 63.7% (58.7–68.4) | 99.6% (97.7–99.9) | 14.4% (9.8–20.6) | 24 (6.0%) | 58.4% |
| **No Hypertension** | 50.0% (18.8–81.2) | 90.7% (85.6–94.1) | 98.2% (94.9–99.4) | 15.0% (5.2–36.0) | 6 (3.2%) | 78.8% |
| **T2D** | 92.9% (68.5–98.7) | 59.5% (51.6–66.9) | 98.9% (94.1–99.8) | 17.3% (10.4–27.4) | 14 (8.4%) | 51.6% |
| **No T2D** | 81.2% (57.0–93.4) | 77.4% (73.1–81.2) | 99.1% (97.3–99.7) | 12.4% (7.4–20.0) | 16 (3.8%) | 69.4% |
| **ASCVD** | 90.9% (72.2–97.5) | 58.0% (51.9–63.8) | 98.7% (95.4–99.6) | 15.3% (10.1–22.4) | 22 (7.7%) | 49.6% |
| **No ASCVD** | 75.0% (40.9–92.9) | 85.5% (81.0–89.0) | 99.2% (97.2–99.8) | 12.2% (5.7–24.2) | 8 (2.6%) | 78.5% |
| **HF** | 94.4% (74.2–99.0) | 44.8% (37.0–53.0) | 98.5% (91.9–99.7) | 17.5% (11.2–26.3) | 18 (11.0%) | 37.0% |
| **No HF** | 75.0% (46.8–91.1) | 82.2% (78.2–85.6) | 99.1% (97.5–99.7) | 10.8% (5.8–19.3) | 12 (2.8%) | 74.1% |

\*Data are presented as point estimate (95% CI), number (percentage), or percentage.
**Abbreviations:** ASCVD, atherosclerotic cardiovascular disease; HF, heart failure; LVH, left ventricular hypertrophy; LVSD, left ventricular systolic dysfunction; NNT, number needed to test; NPV, negative predictive value; PPV, positive predictive value; SHD, structural heart disease; T2D, type 2 diabetes.

### Incremental Value Over KardiaMobile Interpretation

Among the 577 participants with an interpretable native reading, of whom 26 had severe SHD, AI-ECG detected severe SHD with substantially higher sensitivity than the device’s native interpretation (84.6% vs 50.0%; difference, 34.6 percentage points [95% CI, 13.0 to 56.0]), at the cost of lower specificity (73.0% vs 83.3%; difference, -10.3 percentage points [95% CI, -14.3 to -6.4]); PPV was comparable (12.9% vs 12.4%) and NPV modestly higher (99.0% vs 97.2%) (**Table 4**). Reclassification favored AI-ECG (**Figure 2** and **Figure 3**). Among the 472 participants whose tracings were read as normal, 13 had severe SHD, of whom AI-ECG correctly reclassified 10 (76.9%) as high risk; among the 105 read as abnormal, 13 had severe SHD, of whom AI-ECG retained 12 as high risk and misclassified only 1 as low risk. The event NRI was 34.6% and the non-event NRI was -10.3%, yielding a categorical NRI of 24.3% (95% CI, 2.8–45.9) (**Table 5**).

**Figure 2.**
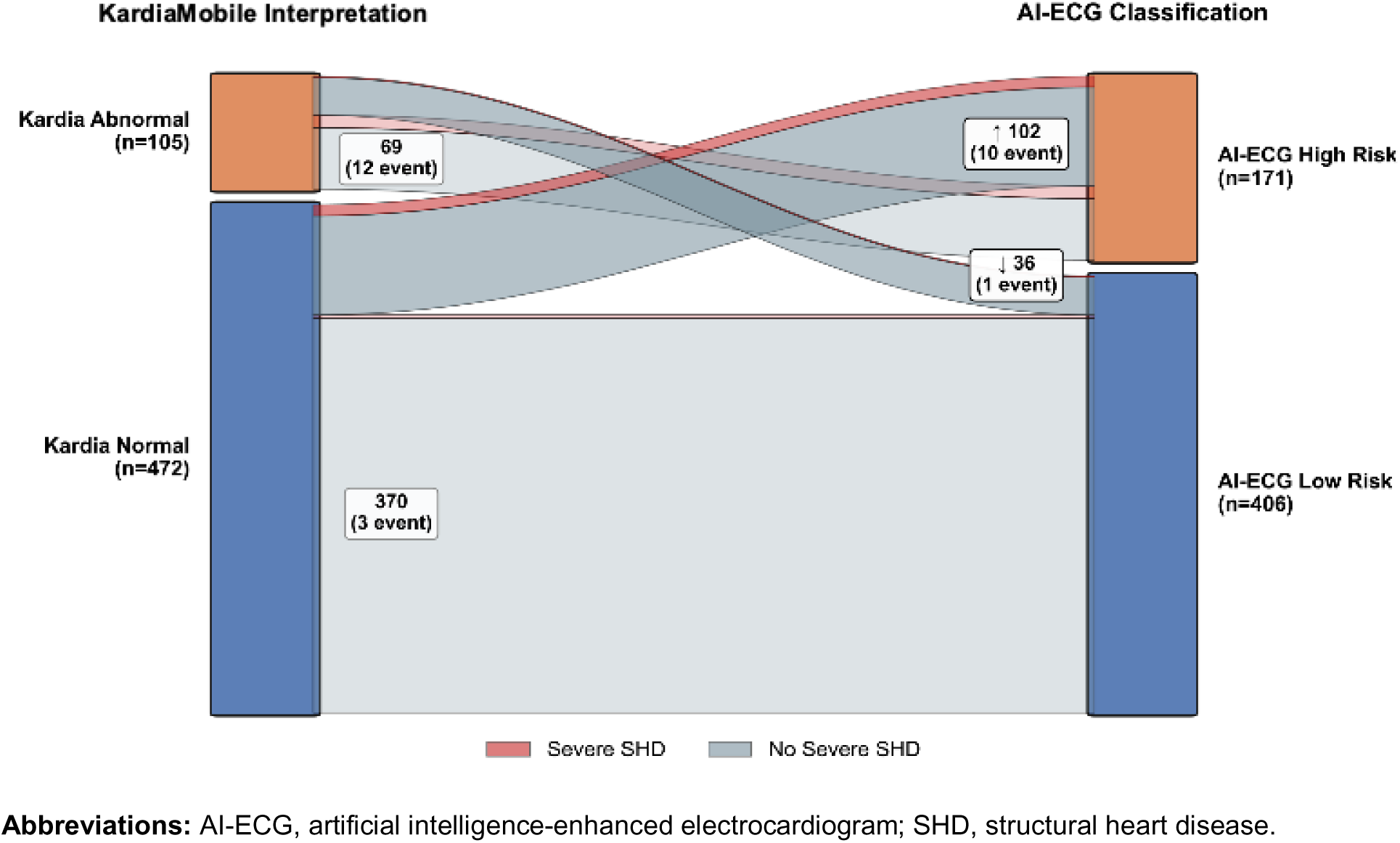
Reclassification from KardiaMobile Interpretation to AI-ECG Risk Stratification for Detecting Severe SHD.

**Figure 3.**
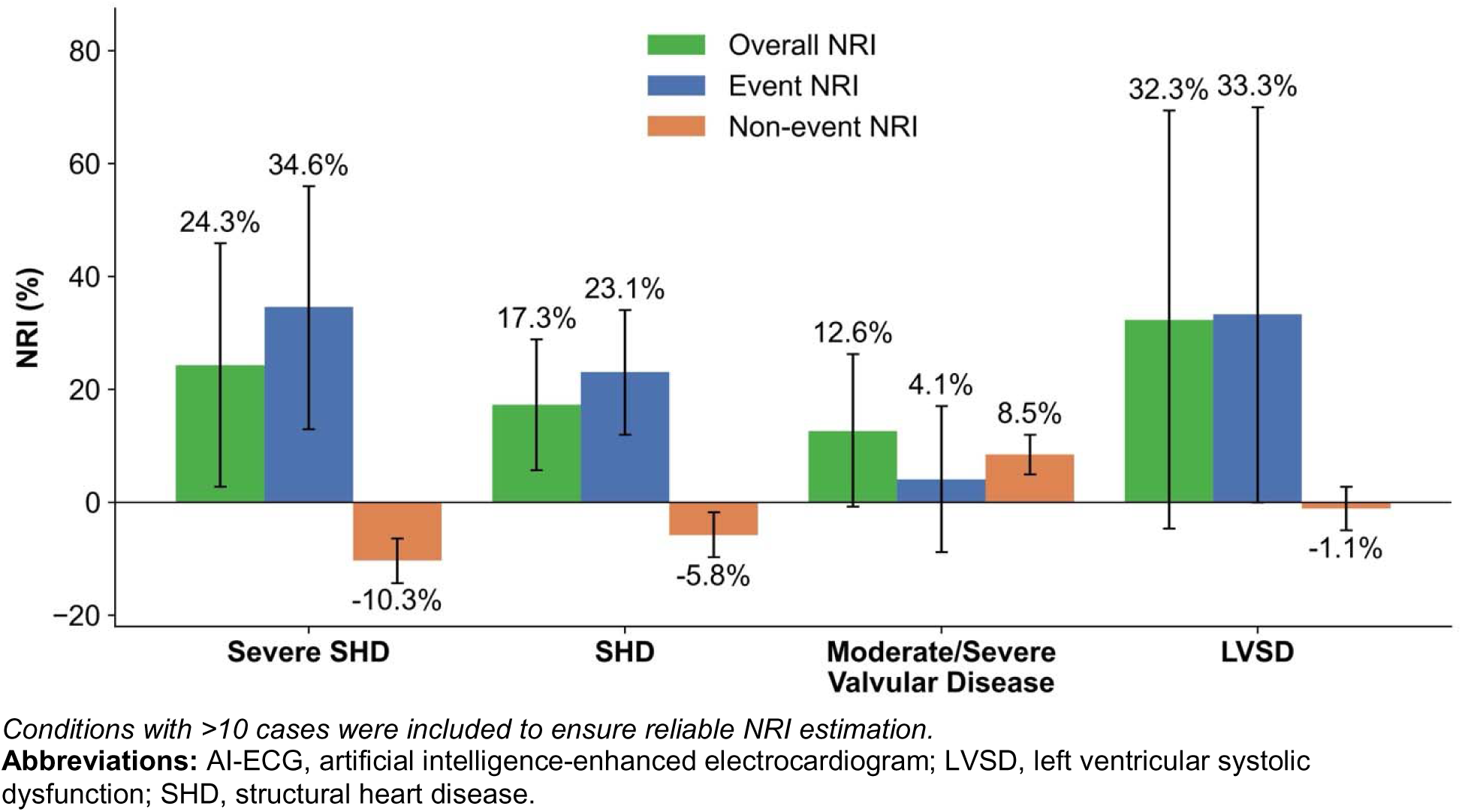
Net Reclassification Improvement of AI-ECG Over KardiaMobile Interpretation for Detecting Structural Heart Disease.

**Table 4.** Head-to-Head Comparison of AI-ECG vs. KardiaMobile Interpretation for Detecting Severe SHD.

| Performance Measure* | AI-ECG | KardiaMobile Interpretation | Difference |
| --- | --- | --- | --- |
| **Sensitivity** | 84.6 (69.6 to 96.6) | 50.0 (30.8 to 69.2) | 34.6 (13.0 to 56.0) |
| **Specificity** | 73.0 (69.1 to 76.6) | 83.3 (80.1 to 86.4) | -10.3 (-14.3 to -6.4) |
| **PPV** | 12.9 (8.1 to 18.1) | 12.4 (6.4 to 19.0) | 0.5 (-4.2 to 5.2) |
| **NPV** | 99.0 (98.0 to 99.8) | 97.2 (95.7 to 98.6) | 1.8 (0.5 to 3.2) |

\*Data are presented as point estimate (95% CI).
**Abbreviations:** AI-ECG, artificial intelligence-enhanced interpretation of electrocardiogram; NPV, negative predictive value; PPV, positive predictive value; SHD, structural heart disease.

**Table 5.** Net Reclassification of Severe SHD Risk: AI-ECG Beyond KardiaMobile Interpretation.

|  | AI-ECG Low Risk | AI-ECG High Risk | Total |
| --- | --- | --- | --- |
| Participants with Severe SHD |  |  |  |
| KardiaMobile Normal | 3 | 10 | 13 |
| KardiaMobile Abnormal | 1 | 12 | 13 |
| Event NRI | 34.6% (95% CI: 13.0 to 56.0) |  |  |
| Participants without Severe SHD |  |  |  |
| KardiaMobile Normal | 367 | 92 | 459 |
| KardiaMobile Abnormal | 35 | 57 | 92 |
| Non-event NRI | -10.3% (95% CI: -14.3 to -6.4) |  |  |
| All Participants |  |  |  |
| KardiaMobile Normal | 370 | 102 | 472 |
| KardiaMobile Abnormal | 36 | 69 | 105 |
| Categorical NRI | 24.3% (95% CI: 2.8 to 45.9) |  |  |

**Abbreviations:** AI-ECG, artificial intelligence-enhanced interpretation of electrocardiogram; CI, confidence interval; NRI, net reclassification improvement; SHD, structural heart disease.

### Number Needed to Test With and Without AI-ECG

Incorporating AI-ECG substantially improved screening efficiency (**Table 2**). For severe SHD, the number needed to test to identify one case decreased from 19.7 under usual care to 6.9 with AI-ECG, a 64.8% reduction. Comparable reductions were observed across the broader composite and individual phenotypes, including 51.3% for SHD, 77.7% for LVSD, 69.5% for severe valvular disease, 65.0% for moderate or severe valvular disease, and 80.9% for severe LVH.

## DISCUSSION

In the ACCESS-SHD study, a noise-adapted 1-lead AI-ECG algorithm detected severe SHD from real-world KardiaMobile recordings with good discrimination (AUROC, 0.872) and provided substantial diagnostic value beyond the rhythm-based interpretation that the device returns to users. Relative to the native interpretation, AI-ECG increased the sensitivity for severe SHD by 34.6 percentage points and yielded a categorical NRI of 24.3%, identifying most cases among individuals reassured by a normal device reading. Incorporating AI-ECG reduced the NNT to detect one case by approximately 65%.

These findings address a question that grows more relevant as portable 1-lead devices reach consumers: whether AI-ECG adds clinically meaningful information beyond the automated interpretation the device already displays. The native interpretation conveys only rhythm and is therefore structurally unable to flag conditions such as LVSD or valvular disease. In our cohort, half of the participants with severe SHD were reassured by a normal device reading. However, applied to the same tracing, AI-ECG reclassified the majority of these individuals as high risk, indicating that the electrical signature of structural disease is recoverable from recordings the device labels as normal. The 13 tracings the device could not interpret were markedly enriched for severe SHD (30.8%), and AI-ECG produced a risk estimate for each, suggesting that the constraints of rule-based rhythm interpretation need not extend to a learned model.

The gain in sensitivity was accompanied by lower specificity and a negative non-event NRI, reflecting the expected trade-off when a more sensitive test is introduced into a screening population. However, because AI-ECG performed better than the device’s native rhythm interpretation, this trade-off did not reduce PPV. This profile is well suited to a front-line triage role, in which a positive AI-ECG result prompts confirmatory echocardiography, a noninvasive and low-risk test. The decision threshold could be tailored to the intended setting, favoring higher specificity where confirmatory imaging capacity is constrained.

Among individual phenotypes, AI-ECG performed best for LVSD (AUROC, 0.943), consistent with prior experience using portable devices.^22–24^ A prospective study employing the 6-lead capability of KardiaMobile 6L device reported an AUROC of 0.924, with 83.4% sensitivity and 88.7% specificity for LVSD detection, among 1635 patients.^24^ Notably, the 1-lead AI-ECG algorithm in ACCESS-SHD detected LVSD comparably well while offering greater acquisition feasibility than 6-lead acquisition. Additionally, our findings extend such single-phenotype evidence to a composite of clinically actionable SHD, which aggregates several low-prevalence conditions to raise event yield and PPV while preserving relevance for downstream testing.^15–18^

Our study has several limitations. First, it was conducted in an echocardiography laboratory among patients already referred for TTE, which likely increased disease prevalence and may have overestimated PPV relative to community screening. Second, this was a single-center study conducted at a large academic hospital in the US. Nonetheless, this design enabled a rigorous prospective comparison against a clinical reference standard and paves the way for future community-based screening efforts. Third, the comparison with the native interpretation was necessarily restricted to tracings the device could interpret, excluding 13 uninterpretable recordings that were enriched for severe SHD. This conservative approach, if anything, understates the advantage of AI-ECG, which produced an estimate for every recording. Fourth, SHD phenotypes were derived from routine clinical TTE interpretation rather than core-laboratory adjudication, and estimates of rarer phenotypes were based on few events. Finally, the native interpretation reflects the device’s current rhythm-classification algorithm, and the downstream clinical impact of AI-ECG-guided screening was not assessed.

## CONCLUSIONS

Applied to real-world 1-lead ECGs from a portable device, AI-ECG meaningfully extended the detection of severe SHD beyond the rhythm-based interpretation available to consumers, identifying clinically actionable disease in individuals reassured by a normal device reading and improving screening efficiency. These findings support portable AI-ECG as a scalable strategy for community-based SHD screening and warrant evaluation in population-based and interventional studies.

## ACKNOWLEDGMENTS

None.

## DATA AVAILABILITY

The data from the Yale New Haven Health System represent protected health information. To protect patient privacy, the Yale Institutional Review Board does not allow the sharing of these data.

## REFERENCES

1. Wolfe NK, Mitchell JD, Brown DL. The independent reduction in mortality associated with guideline-directed medical therapy in patients with coronary artery disease and heart failure with reduced ejection fraction. Eur Heart J Qual Care Clin Outcomes 2021;7:416–421.

2. Olivotto I, Oreziak A, Barriales-Villa R, Abraham TP, Masri A, Garcia-Pavia P, Saberi S, Lakdawala NK, Wheeler MT, Owens A, Kubanek M, Wojakowski W, Jensen MK, Gimeno-Blanes J, Afshar K, Myers J, Hegde SM, Solomon SD, Sehnert AJ, Zhang D, Li W, Bhattacharya M, Edelberg JM, Waldman CB, Lester SJ, Wang A, Ho CY, Jacoby D, EXPLORER-HCM study investigators. Mavacamten for treatment of symptomatic obstructive hypertrophic cardiomyopathy (EXPLORER-HCM): a randomised, double-blind, placebo-controlled, phase 3 trial. Lancet 2020;396:759–769.

3. Sara JD, Toya T, Taher R, Lerman A, Gersh B, Anavekar NS. Asymptomatic left ventricle systolic dysfunction. Eur Cardiol 2020;15:e13.

4. Manning WJ. Asymptomatic aortic stenosis in the elderly: a clinical review. JAMA 2013;310:1490–1497.

5. Wang TJ, Evans JC, Benjamin EJ, Levy D, LeRoy EC, Vasan RS. Natural history of asymptomatic left ventricular systolic dysfunction in the community. Circulation 2003;108:977–982.

6. Wang TJ, Levy D, Benjamin EJ, Vasan RS. The Epidemiology of Asymptomatic Left Ventricular Systolic Dysfunction: Implications for Screening. Ann Intern Med 2003;138:907– 916.

7. Arcy JL d’, Coffey S, Loudon MA, Kennedy A, Pearson-Stuttard J, Birks J, Frangou E, Farmer AJ, Mant D, Wilson J, Myerson SG, Prendergast BD. Large-scale community echocardiographic screening reveals a major burden of undiagnosed valvular heart disease in older people: the OxVALVE Population Cohort Study. Eur Heart J 2016;37:3515–3522.

8. Maron MS, Hellawell JL, Lucove JC, Farzaneh-Far R, Olivotto I. Occurrence of clinically diagnosed hypertrophic cardiomyopathy in the United States. Am J Cardiol 2016;117:1651– 1654.

9. Motazedian P, Prosperi-Porta G, Hibbert B, Jalal H, Labinaz M, Burwash IG, Abdel-Razek O, Di Santo P, Simard T, Wells G, Coyle D. Cost-effectiveness of population screening for aortic stenosis. Eur Heart J Qual Care Clin Outcomes 2025;11:378–387.

10. Galasko GI, Barnes SC, Collinson P, Lahiri A, Senior R. What is the most cost-effective strategy to screen for left ventricular systolic dysfunction: natriuretic peptides, the electrocardiogram, hand-held echocardiography, traditional echocardiography, or their combination? Eur Heart J 2006;27:193–200.

11. Steinberg DH, Staubach S, Franke J, Sievert H. Defining structural heart disease in the adult patient: current scope, inherent challenges and future directions. Eur Heart J Suppl 2010;12:E2–E9.

12. Aminorroaya A, Biswas D, Pedroso AF, Khera R. Harnessing artificial intelligence for innovation in interventional cardiovascular care. J Soc Cardiovasc Angiogr Interv 2025;4:102562.

13. Oikonomou EK, Khera R. Artificial intelligence-enhanced patient evaluation: bridging art and science. Eur Heart J 2024;45:3204–3218.

14. Dhingra LS, Croon PM, Batinica B, Aminorroaya A, Pedroso AF, Khera R. Artificial intelligence-enhanced electrocardiography for heart failure screening and risk stratification. Curr Heart Fail Rep 2026;23.

15. Aminorroaya A, Dhingra LS, Pedroso AF, Shankar SV, Coppi A, Khunte A, Foppa M, Brant LCC, Barreto SM, Ribeiro ALP, Krumholz HM, Oikonomou EK, Khera R. Development and multinational validation of an ensemble deep learning algorithm for detecting and predicting structural heart disease using noisy single-lead electrocardiograms. Eur Heart J Digit Health 2025.

16. Dhingra LS, Aminorroaya A, Sangha V, Pedroso AF, Shankar SV, Coppi A, Foppa M, Brant LCC, Barreto SM, Ribeiro ALP, Krumholz HM, Oikonomou EK, Khera R. Ensemble deep learning algorithm for structural heart disease screening using electrocardiographic images: PRESENT SHD. J Am Coll Cardiol 2025;85:1302–1313.

17. Ulloa-Cerna AE, Jing L, Pfeifer JM, Raghunath S, Ruhl JA, Rocha DB, Leader JB, Zimmerman N, Lee G, Steinhubl SR, Good CW, Haggerty CM, Fornwalt BK, Chen R. rECHOmmend: An ECG-Based Machine Learning Approach for Identifying Patients at Increased Risk of Undiagnosed Structural Heart Disease Detectable by Echocardiography. Circulation.

18. Poterucha TJ, Jing L, Ricart RP, Adjei-Mosi M, Finer J, Hartzel D, Kelsey C, Long A, Rocha D, Ruhl JA, vanMaanen D, Probst MA, Daniels B, Joshi SD, Tastet O, Corbin D, Avram R, Barrios JP, Tison GH, Chiu I-M, Ouyang D, Volodarskiy A, Castillo M, Roedan Oliver FA, Malta PP, Ye S, Rosner GF, Dizon JM, Ali SR, Liu Q, Bradley CK, Vaishnava P, Waksmonski CA, DeFilippis EM, Agarwal V, Lebehn M, Kampaktsis PN, Shames S, Beecy AN, Kumaraiah D, Homma S, Schwartz A, Hahn RT, Leon M, Einstein AJ, Maurer MS, Hartman HS, Hughes JW, Haggerty CM, Elias P. Detecting structural heart disease from electrocardiograms using AI. Nature 2025;644:221–230.

19. Petek BJ, Al-Alusi MA, Moulson N, Grant AJ, Besson C, Guseh JS, Wasfy MM, Gremeaux V, Churchill TW, Baggish AL. Consumer wearable health and fitness technology in cardiovascular medicine: JACC state-of-the-art review. J Am Coll Cardiol 2023;82:245–264.

20. Spatz ES, Ginsburg GS, Rumsfeld JS, Turakhia MP. Wearable digital health technologies for monitoring in cardiovascular medicine. N Engl J Med 2024;390:346–356.

21. Khunte A, Sangha V, Oikonomou EK, Dhingra LS, Aminorroaya A, Mortazavi BJ, Coppi A, Brandt CA, Krumholz HM, Khera R. Detection of left ventricular systolic dysfunction from single-lead electrocardiography adapted for portable and wearable devices. npj Digital Medicine 2023;6:1–10.

22. Attia ZI, Harmon DM, Dugan J, Manka L, Lopez-Jimenez F, Lerman A, Siontis KC, Noseworthy PA, Yao X, Klavetter EW, Halamka JD, Asirvatham SJ, Khan R, Carter RE, Leibovich BC, Friedman PA. Prospective evaluation of smartwatch-enabled detection of left ventricular dysfunction. Nat Med 2022;28:2497–2503.

23. Bachtiger P, Petri CF, Scott FE, Ri Park S, Kelshiker MA, Sahemey HK, Dumea B, Alquero R, Padam PS, Hatrick IR, Ali A, Ribeiro M, Cheung W-S, Bual N, Rana B, Shun-Shin M, Kramer DB, Fragoyannis A, Keene D, Plymen CM, Peters NS. Point-of-care screening for heart failure with reduced ejection fraction using artificial intelligence during ECG-enabled stethoscope examination in London, UK: a prospective, observational, multicentre study. The Lancet Digital Health 2022;4:e117–e125.

24. Lim J, Lee HS, Han GI, Kang S, Jang J-H, Jo Y-Y, Son JM, Lee MS, Kwon J-M, Lee S-P. Artificial intelligence-enhanced six-lead portable electrocardiogram device for detecting left ventricular systolic dysfunction: a prospective single-centre cohort study. European Heart Journal - Digital Health 2025;6:476–485.

25. FDA. 510(k) Premarket Notification for Kardiamobile 6L https://www.accessdata.fda.gov/cdrh_docs/pdf22/K220350.pdf

26. FDA. 510(k) Premarket Notification for Apple Watch ECG App https://www.accessdata.fda.gov/cdrh_docs/pdf20/K201525.pdf

27. Mannhart D, Lischer M, Knecht S, Fay de Lavallaz J du, Strebel I, Serban T, Vögeli D, Schaer B, Osswald S, Mueller C, Kühne M, Sticherling C, Badertscher P. Clinical Validation of 5 Direct-to-Consumer Wearable Smart Devices to Detect Atrial Fibrillation: BASEL Wearable Study. JACC Clin Electrophysiol 2023;9:232–242.

28. Mitchell C, Rahko PS, Blauwet LA, Canaday B, Finstuen JA, Foster MC, Horton K, Ogunyankin KO, Palma RA, Velazquez EJ. Guidelines for performing a comprehensive transthoracic echocardiographic examination in adults: Recommendations from the American society of echocardiography. J Am Soc Echocardiogr 2019;32:1–64.

29. Lang RM, Badano LP, Mor-Avi V, Afilalo J, Armstrong A, Ernande L, Flachskampf FA, Foster E, Goldstein SA, Kuznetsova T, Lancellotti P, Muraru D, Picard MH, Rietzschel ER, Rudski L, Spencer KT, Tsang W, Voigt J-U. Recommendations for cardiac chamber quantification by echocardiography in adults: an update from the American Society of Echocardiography and the European Association of Cardiovascular Imaging. J Am Soc Echocardiogr 2015;28:1–39.e14.

30. Aminorroaya A, Dhingra LS, Oikonomou EK, Khera R. Evaluation of a machine learning-guided strategy for elevated Lipoprotein(a) screening in health systems. Circ Genom Precis Med 2025;18:e004632.

31. Aminorroaya A, Dhingra LS, Oikonomou EK, Saadatagah S, Thangaraj P, Vasisht Shankar S, Spatz ES, Khera R. Development and multinational validation of an algorithmic strategy for high Lp(a) screening. Nature Cardiovascular Research 2024;3:558–566.

32. Dhingra LS, Pedroso AF, Aminorroaya A, Rajpura J, Mehanna S, Tonnu-Mihara I, Khera R. A real-world evaluation of longitudinal healthcare expenses in a health system registry of type-2 diabetes mellitus and cardiovascular disease enabled by the 21st century cures act. Am J Prev Cardiol 2026;25:101425.

33. Vasisht Shankar S, Oikonomou EK, Khera R. CarDS-Plus ECG platform. arXiv.

34. Wong KC, Klimis H, Lowres N, Huben A von, Marschner S, Chow CK. Diagnostic accuracy of handheld electrocardiogram devices in detecting atrial fibrillation in adults in community versus hospital settings: a systematic review and meta-analysis. Heart 2020;106:1211– 1217.

35. Halcox JPJ, Wareham K, Cardew A, Gilmore M, Barry JP, Phillips C, Gravenor MB. Assessment of remote heart rhythm sampling using the AliveCor heart monitor to screen for atrial fibrillation: The REHEARSE-AF study. Circulation 2017;136:1784–1794.

36. Wilson EB. Probable Inference, the Law of Succession, and Statistical Inference. J Am Stat Assoc 1927;22:209–212.

37. Pencina MJ, D’Agostino RB Sr, D’Agostino RB Jr, Vasan RS. Evaluating the added predictive ability of a new marker: from area under the ROC curve to reclassification and beyond. Stat Med 2008;27:157–172; discussion 207-12.

